# Quantum Neural Network Tuning and Performance Evaluation for a Breast Cancer Dataset

**DOI:** 10.1101/2025.10.03.25336905

**Authors:** Seung Joo Lee, Thomas JS Durant, Sarah Dudgeon, Brent Nelson, H. Patrick Young, Gustav Horn, Wade L. Schulz

**Author notes:** Corresponding Author: 55 Park St, New Haven, CT 06510.

## Abstract

Model tuning with the optimization of pipeline configuration is a well-established practice for the development of machine learning models. However, this often entails an exhaustive search process, especially as the parameter space expands with increasing model complexity. In the emerging field of quantum machine learning (QML), there is limited literature on the effects of configuration parameters, especially quantum-specific ones, and their choices on model performance. To address this gap, here we present a study exploring the impacts of data scaling and configuration parameters in quantum neural network (QNN) development using beta regression. Our experiments with two benchmark datasets showed that a well-tuned QNN can achieve predictive performance comparable to its classical counterparts. Our findings also demonstrate useful reference points of QNN model tuning to support a more efficient parameter optimization process.

## INTRODUCTION

Artificial intelligence and machine learning (AI/ML) are widely used for many biomedical and healthcare applications, ranging from basic research to clinical care. Literature has shown promise of AI/ML applications in health sector with empirical evidence in areas including clinical decision support, clinical predictive health and health monitoring, public health applications, and improving health services delivery and data fusion^1,2,3^. However, training these models is computationally intensive and often requires large, well-labeled datasets^4^. Quantum machine learning (QML), an emerging subfield of quantum computing, has t^1^he potential to increase the efficiency and/or accuracy of AI/ML algorithms in biomedicine^1^. Quantum computers, which rely on quantum bits, or qubits, provide a fundamentally new approach to computing. AI/ML algorithms built on these systems can leverage the quantum properties of superposition and entanglement to provide several possible advantages over classical computing, including shorter training times, increased performance, and the ability to train models on smaller datasets^5^.

While classical AI/ML algorithms cannot be directly implemented on quantum computers, the increased accessibility of quantum computers has allowed algorithm developers to implement quantum-enabled AI/ML algorithms. These include classical AI/ML equivalents, such as variational quantum classifiers (VQC) and quantum neural networks (QNN), along with quantum-specific ML algorithms such as quantum phase estimation (QPE) and quantum circuit learning (QCL). Studies used quantum-enabled AI/ML algorithms that leverage quantum computing to analyze complex healthcare and biomedical datasets^1,6–8^. For example, VQCs have been used to predict the presence of diseases such as breast cancer, heart disease, and diabetes^7^. Another study compared the predictive performance of three quantum ML algorithms, a quantum state vector classifier (QSVC), a QNN, and a VQC, with classical ML classifiers using the UCI heart disease dataset and reported the QSVC model outperformed other models with an accuracy of 90.16%^8^.

Configuration parameters are parameters that need to be set in advance of the model training process. Classical AI/ML development alike, quantum AI/ML development also involves data preprocessing and configuration parameters tuning^9,10^. Data preprocessing and configuration parameters tuning are crucial steps in the AI/ML model development and lifecycle^11^. Data preprocessing involves techniques such as data cleansing, feature engineering, and scaling to ensure the quality and usability of data that can impact downstream algorithm performance ^12–16^. For quantum neural networks (QNN), there are quantum-specific configuration parameters such as quantum encoding method, circuit design, and transpiler optimizer that can be tuned and optimized to improve model performance. Grid search is one of the most commonly used search methods for tuning AI/ML models and optimizing pipeline configurations to improve model performance^17,18^. Since it covers a complete set of all pre-train parameters, it often entails a resource-intensive and time-consuming process as each additional parameter adds to the number of runs and search time^19^. To address this challenge, studies have been conducted to develop general guidance for default parameter settings to help scope the configuration parameter space and make the pipeline configuration process more efficient^20^.

Given the recency of the field and the unique architectures of quantum AI/ML, few studies have discussed their configuration pipelines, model optimization, and the effect of these choices on downstream model performance^9,10^. Moussa et al. (2024)^9^, for example, used functional ANOVA to identify important hyperparameters in QNN development. The results of the study revealed the learning rate to be the most important hypermeter followed by the type of entangling gate. Zaman et al. (2024)^10^ conducted a correlation analysis of hyperparameters that configure quantum circuit design in the context of hybrid QNN. The study identified the number of shots as a key hyperparameter directly proportional to the model accuracy. In the same study, moreover, the hybrid QNN with basic entangling gates and the one with strongly entangling gates demonstrated very similar performance in terms of accuracy, which was partially in line with the findings from Moussa et al. (2024). However, the literature has only partially addressed the vast range of quantum-specific configuration parameters, individual impacts of their configuration values, and their interdependencies, leaving gaps in understanding their impacts on QNN model performance.

In this manuscript, we evaluate the effects of data preprocessing methods and model configuration parameters on the predictive performance of a QNN model using a benchmark biomedical dataset and a synthetic dataset, thereby addressing the existing gaps in quantum AI/ML model tuning and parameters optimization (Supplementary Figure 1). Additionally, we compared QNN against its classical ML counterpart, Multi-Layer Perceptron (MLP), to examine its practical applicability in biomedical and healthcare research, given current limitations of quantum computing.

## METHODS

### Datasets and Data Preprocessing

Two datasets, the Wisconsin Diagnostic Breast Cancer (WDBC) dataset and a synthetic dataset, were used in this study. The WDBC dataset is a benchmark dataset comprised of a binary target variable and 30 continuous features computed from a digitized image of a fine needle aspirate (FNA) of a breast mass ^21^. These features describe the characteristics of nuclei images and are used to classify tumors as either malignant or benign. From the total sample of 569 observations, we extracted the target variable and the five most important features identified using the Random Forest feature selection method. These features are *concave_points, radius_worst, perimeter_worst, area_worst*, and *concave_points_worst*. The Random Forest feature selection was performed in Python (version 3.9) using the Scikit-Learn library (version 1.5.1).

The synthetic dataset consists of 569 randomly generated nonlinear data points. Initially, we used the *make_classification* function from Scikit-Learn in Python to generate a binary random classification dataset with 1,000 samples and five feature variables. We set the number of informative features to three (*n_informative*=3), the number of redundant features to two *(n_redundant*=2), and the number of clusters per class to three *(n_clusters_per_class=*3). A non-linear transformation was then to the data values generated. To create the final dataset, we first randomly sampled 212 positive (y=1) and then 357 negative (y=0) samples from the initial dataset, aligning the class balance with that of the breast cancer dataset.

Both datasets were scaled in the data preprocessing stage, using *StandardScaler* (standard) and the *MinMaxScaler* with two different ranges, 0 to 1 and 0 to 2*p. We used 80/20 80/20 train and test split ratio for all models.

### Classical Neural Network

We implemented a multi-layer perceptron (MLP) classifier with Scikit-Learn to determine the baseline performance of a classical neural network. Our MLP model consisted of two hidden layers each with five and two neurons, respectively, and employed the L-BFGS optimizer and the ReLU activation function. We did not implement configuration parameter tuning for MLP and applied mostly the default settings available in the library. All programming for classical neural network development was implemented in Python using Scikit-Learn.

### Quantum Neural Network

We implemented our quantum neural network models in Python with IBM’s Qiskit SDK (version 1.1.0) ^22^ and Scikit-Learn. Our quantum models were implemented based on the Sampler primitive with FakeBelem, a 5-qubit backend configuration available in the Qiskit library. Qiskit’s Aer Simulator backend with the statevector simulation method was implemented to run noisy quantum circuits and ZZFeatureMap, a feature map method with second-order Pauli-Z evolution circuit, was used to encode the classical binary data into the quantum state. The entanglement structure was set to ‘full’, where each qubit is entangled with all the others. We experimented with two types of quantum simulator, one noisy and another ideal. A noisy simulator replicated the actual noise profile of quantum system while an ideal simulator reflected a perfect computing system without noise. We performed the configuration parameter tuning and model optimization with the ansatz circuit type, the number of ansatz repetitions, optimizer, optimization level, input gradients, and the number of feature map repetitions (Table 1, Fig. 1).

**Table 1.** Configuration parameters and optimization search space.

| Parameters | Values | Description |
| --- | --- | --- |
| Data Scaler | [Standard, Min0-Max1, Min0-Max$2\pi$] | Data scaling is a data preprocessing technique that standardizes features to a common scale, preventing large-valued features from dominating model training and ensuring equal contribution across variables. This improves model performance, stability, and accuracy, and is particularly important for distance-based algorithms such as neural networks. |
| Feature Map Reps | [1, 3, 5] | Quantum feature map is an essential data preprocessing step needed to make the data in classical computing bits compatible with a quantum environment. It nonlinearly maps a datum from classical computing bits to the quantum state space (i.e., the Hilbert space of density matrices) by using a quantum circuit. We employed the ZZFeatureMap that encodes not only the features but also their second-order interactions (i.e., entanglement) in accordance with the connectivity graph and the classical data map. Feature Map Repetitions identifies the number of times to repeat the feature map circuit and control for its circuit depth. More feature map reps can result in the quantum feature map with higher dimensionality and thus complexity. |
| Ansatz | [EfficientSU2, RealAmplitudes] | In quantum computing, ansatz is a basic architecture of a circuit or a set of gates that acts as a starting point for model optimization. We included ansatz as part of our configuration parameters space because the fit of this initial circuit to the model and task can impact the efficiency and accuracy of the quantum algorithm. EfficientSU2 is a parameterized circuit architecture which consists of qubits with multiple layers of single qubit operations and their entanglement. Real Amplitudes is structured with all qubits initialized with single rotations and their entanglement. EfficientSU2 is often considered more versatile and expressive due to its architecture with multiple rotation axes and denser entanglement, suitable for problems with more complex patterns and/or requiring higher entanglement. |
| Ansatz Reps | [1, 3, 5] | Feature map repetitions alike, the ansatz repetitions set the number of times to repeat the ansatz circuit. Increasing this model parameter adds depth to the ansatz circuit, affecting its expressiveness and capability to understand and interpret complex data patterns. |
| Optimization Level | [0, 1, 3] | In Qiskit, the optimization level ranges from 0 to 3. When at 0, there is no optimization. Higher optimization levels |
|  |  | indicate more optimized circuits to account for the noise profile during transpilation time and computational effort needed. |
| Optimizer | [COBYLA, SPSA] | We compared two widely adopted classical local optimizers, Constrained Optimization by Linear Approximation (COBYLA) and Simultaneous Perturbation Stochastic Approximation (SPSA) for circuit optimization with the transpilation process. The performance of optimizers is contingent on various quantum environment factors, including gradients and noise. COBYLA is a derivative-free constrained optimization method that takes the objective and constraint values when the derivative of the objective function is unknown. For each iteration, it solves a linear programming problem and updates the linear approximations for the objective and constraints to decrease the radius of a “trust region”* and reach a constrained optimum. SPSA uses objective function measurements from “gradient approximation” through perturbations. Because it only takes two objective function measurements per iteration regardless of the dimensions and scale of the problem, it is perceived to be highly efficient compared to the optimizers taking direct measurements of gradients, especially for large-scale problems or simulation optimization.<br>*“trust region”: the subset of the region of the objective function that is approximated by a model function |
| Input Gradients | [True, False] | Gradient backpropagation algorithm when training QNN. We used Qiskit’s backend sampler to compute the gradients. |

**Figure 1.**
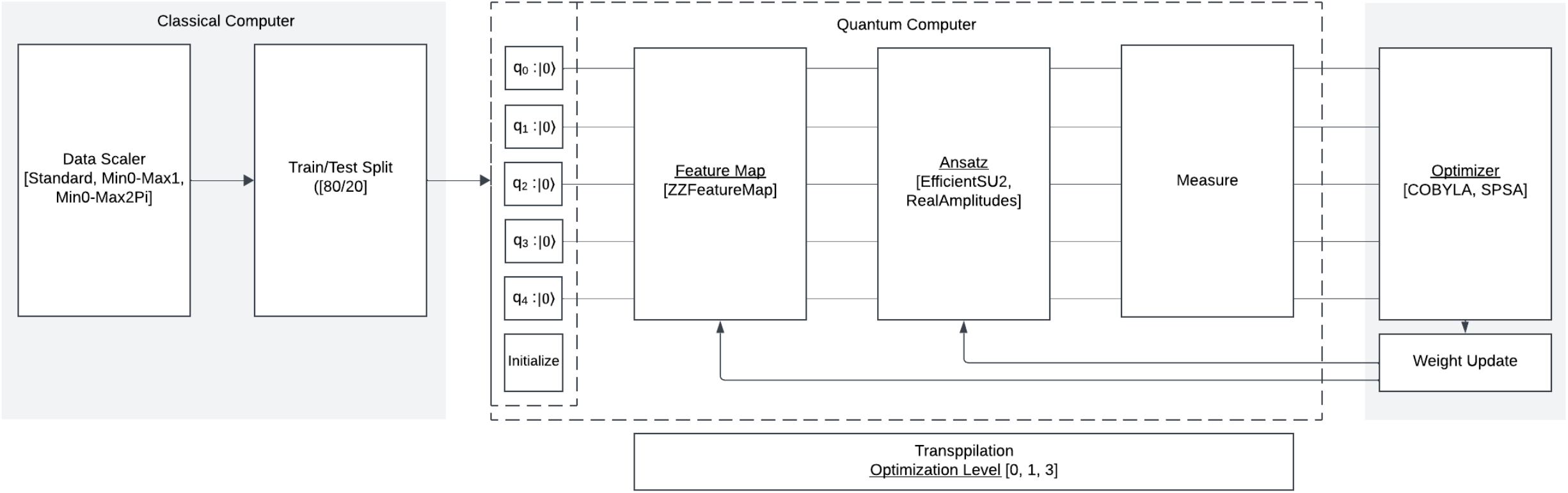
Architectural diagram of the QNN highlighting classical and quantum computing components.

### Performance Metrics and Model Performance Data

Performance metrics were obtained from each optimization experiment to evaluate the predictive performance of both the classical and quantum neural network models with the consistent holdout datasets. We used evaluations functions available in Scikit-Learn, F1 score, precision, accuracy, and recall. For the QNN models, we performed a grid search of data scalers and configuration parameters, resulting in a total of 648 runs for each data set on both an ideal and noisy simulator. We repeated each grid search three times, which amounted to a total of 1,944 runs per QNN model, to address the variability in performance results. For the classical MLP models, we evaluated the impact of different data scaling techniques to assess for potential variability. All experiments were logged using the Weights & Biases (W&B) wandb python library (version 0.17.8).

### QNN Configuration parameters Evaluation Using Beta Regression

We used beta regression to conduct a statistical analysis of the impact of the configuration parameters on the predictive performance of the QNN models. Beta regression is a type of regression tailored for contexts where the response variable (*y*) is measured continuously on the standard interval, 0 < *y* < 1, and is assumed to be beta distributed with a standard parameterization using two shape parameters, *p* and *q*^23^. The density of the beta distribution is defined as^23,24^:

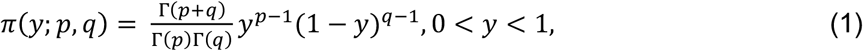

where *p* > 0, *q* > 0 and Γ(·) is the gamma function. The estimated mean and variance of, respectively, are:

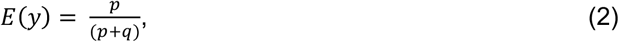

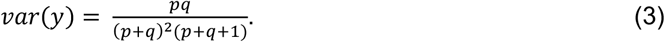

We chose beta regression to fit our model performance data for the following reasons. Firstly, beta regression is useful for situations where the target variable is continuous and within the range of (0, 1) and is assumed to have meaningful associations with the feature variables^23,25^. Secondly, the model has shape parameters *p* and *q* to calculate the main parameters, the mean (m) and the precision parameter (j), that calibrate the shape and density of the distribution curve. This approach naturally incorporates data features such as skewness and reserves assumptions (e.g., homoscedasticity) held on other comparable Generalized Linear Models (GLMs) such as Ordinary Least Squares (OLS) linear regression and logistic regression^25^. Lastly, interpreting the parameters of beta regression with logit link is relatively straightforward and intuitive as of an odds ratio compared to that of a linear regression with transformed target variables^23^. We regressed the F1 Score on the data scalers and six model configuration parameters. A p-value threshold of <0.05 was used to determine the statistical significance of the coefficients. All data cleansing/processing, statistical modeling, and analysis were conducted in Python with Scikit-Learn, the Pandas library (version 2.2.2) and the Statsmodels library (version 0.14.1).

### Data and Code Availability

Data are available from the original publication^21^. Code was written and independently reviewed by two separate authors and is available at https://github.com/acomphealth/quantum-neural-network.git.

## RESULTS

### Configuration choices dramatically impact QNN’s model performance

The predictive performance of all QNN models varied considerably by the selection of data scaling methods and configuration parameter values (Fig. 2). For the breast cancer dataset, the F1 score ranged from 0.12 to 0.89 using a noisy simulator (mean = 0.52, standard deviation (SD) = 0.12), and from 0.18 to 0.92 using an ideal simulator (mean = 0.60, SD = 0.11). For the synthetic dataset using a noisy simulator, the mean F1 score was 0.33 (SD = 0.15), ranging from 0.01 to 0.78, and using an ideal simulator, it was 0.48 (SD = 0.10) and ranged from 0.04 and 0.82. The variability in the distributions of performance metrics indicates that QNN model performance can be affected by data scaling methods and the tuning of model configuration parameters (Table 2).

**Table 2.**
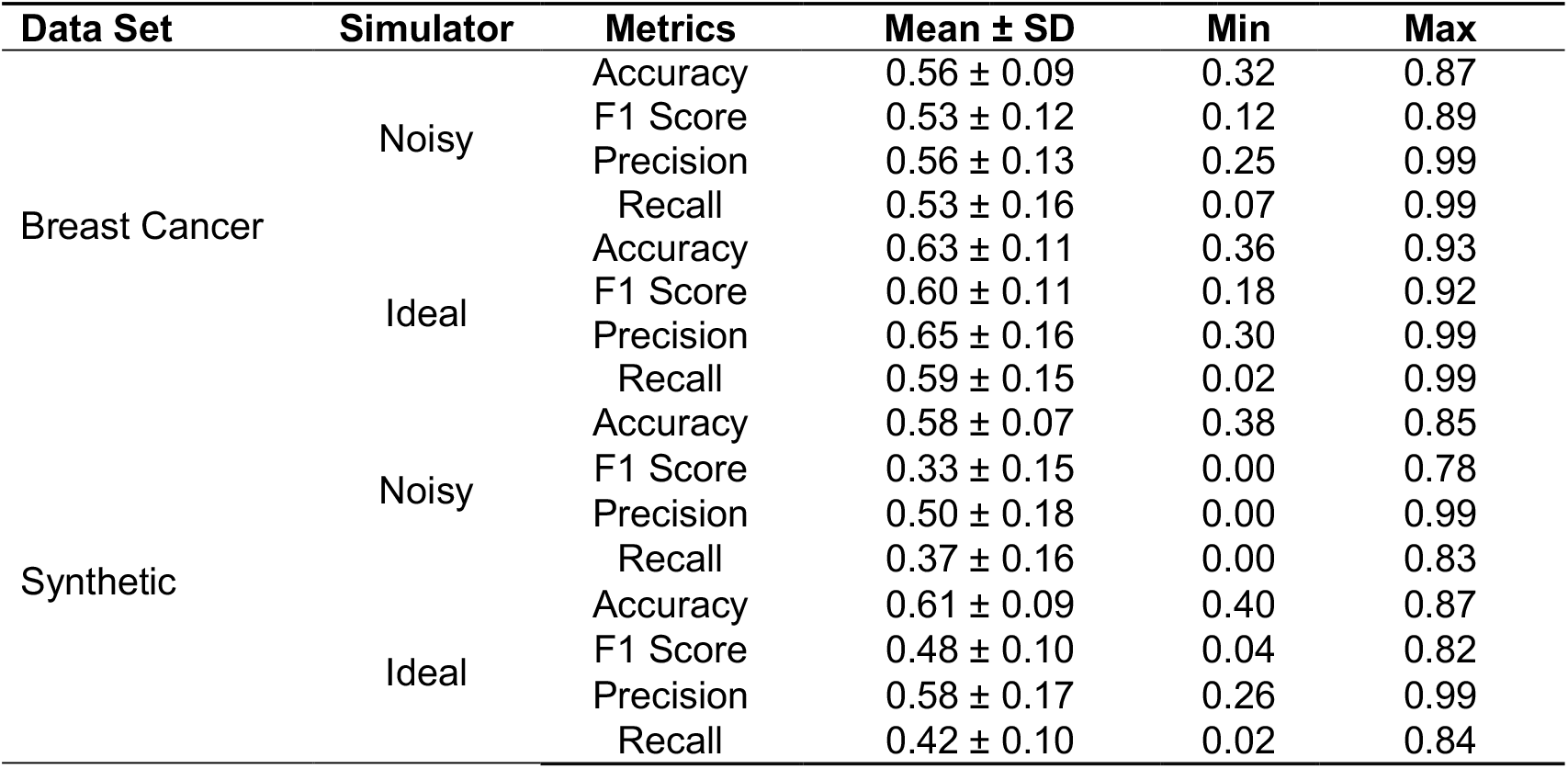
Descriptive statistics of QNN’s predictive performance.

| **Data Set** | **Simulator** | **Metrics** | **Mean ± SD** | **Min** | **Max** |
| --- | --- | --- | --- | --- | --- |
| Breast Cancer | Noisy | Accuracy | 0.56 ± 0.09 | 0.32 | 0.87 |
| | | F1 Score | 0.53 ± 0.12 | 0.12 | 0.89 |
| | | Precision | 0.56 ± 0.13 | 0.25 | 0.99 |
| | | Recall | 0.53 ± 0.16 | 0.07 | 0.99 |
| | Ideal | Accuracy | 0.63 ± 0.11 | 0.36 | 0.93 |
| | | F1 Score | 0.60 ± 0.11 | 0.18 | 0.92 |
| | | Precision | 0.65 ± 0.16 | 0.30 | 0.99 |
| | | Recall | 0.59 ± 0.15 | 0.02 | 0.99 |
| Synthetic | Noisy | Accuracy | 0.58 ± 0.07 | 0.38 | 0.85 |
| | | F1 Score | 0.33 ± 0.15 | 0.00 | 0.78 |
| | | Precision | 0.50 ± 0.18 | 0.00 | 0.99 |
| | | Recall | 0.37 ± 0.16 | 0.00 | 0.83 |
| | Ideal | Accuracy | 0.61 ± 0.09 | 0.40 | 0.87 |
| | | F1 Score | 0.48 ± 0.10 | 0.04 | 0.82 |
| | | Precision | 0.58 ± 0.17 | 0.26 | 0.99 |
| | | Recall | 0.42 ± 0.10 | 0.02 | 0.84 |

**Figure 2.**
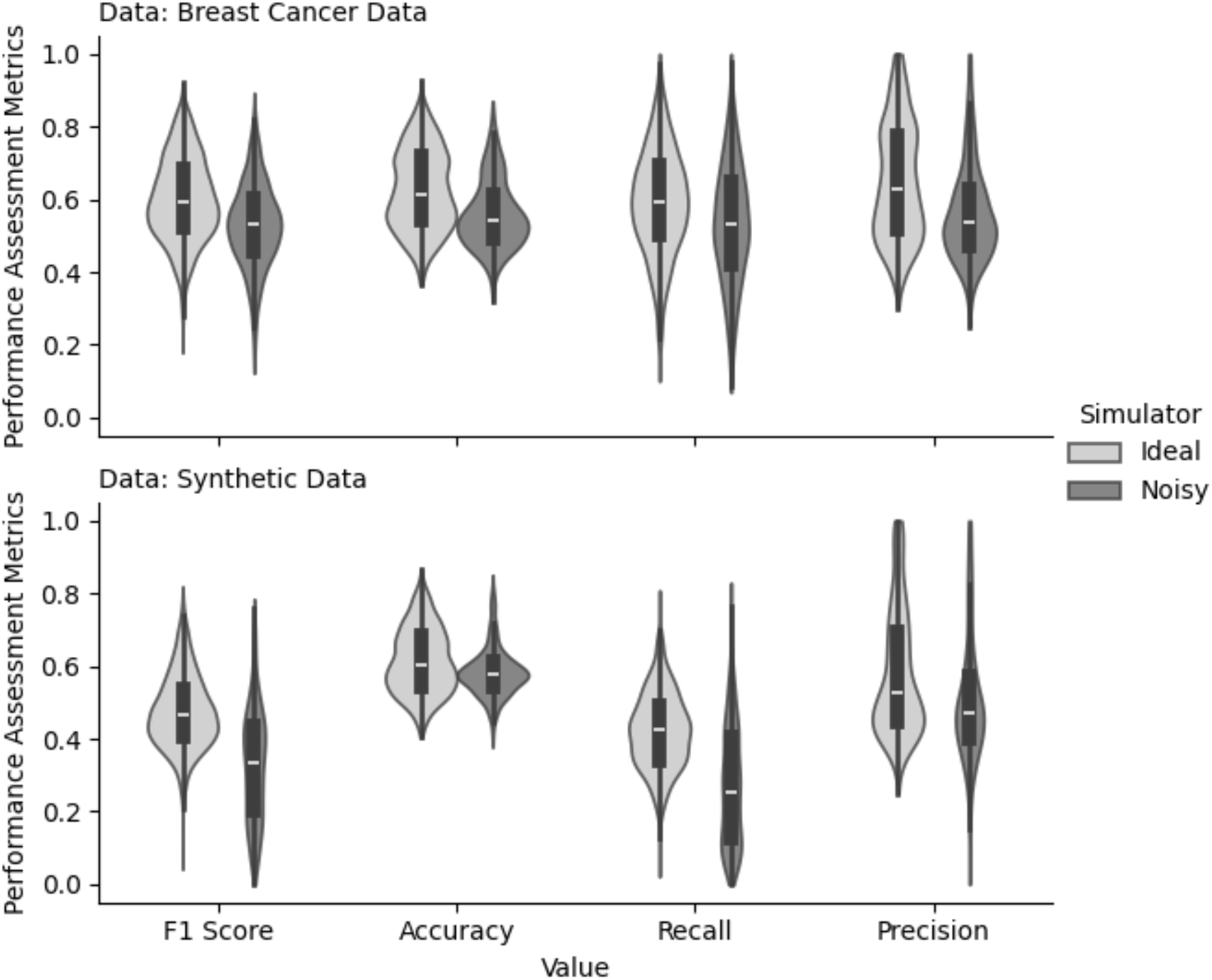
QNN performance results by data set and simulator type. For each QNN model, the predictive performance was measured on four different metrics, F1 score, accuracy, recall, and precision.

### Multiple configuration choices demonstrated consistent impacts on downstream classification performance

We used beta regression to analyze the impacts of data scaling and configuration parameters on QNN model performance statistically. Several key impacts were consistent across all QNN models controlling for their pairwise interaction effects (Table 3, Fig. 3). For example, data scaling with the standard scaler resulted in the lowest performance scores for all models, holding other parameter values constant. For the breast cancer dataset using a noisy simulator, the coefficient of standardizing the data compared to scaling to a range of 0 and 1 was −0.22 ± 0.07 and using an ideal simulator was −0.21 ± 0.06. For the synthetic dataset, likewise, the QNN model’s predictive performance worsened when standardizing the data compared to scaling between 0 and 1, by −0.29 ± 0.02 using a noisy simulator and by −0.22 ± 0.00 using an ideal simulator. Increasing the number of feature map repetitions also had statistically significant negative effects on F1 scores in all QNN models. For the breast cancer dataset with a noisy simulator, the corresponding coefficient was −0.13 ± 0.01 and with an ideal simulator was −0.02 ± 0.01. For the synthetic dataset with a noisy simulator, the coefficient was - 0.44 ± 0.03 and −0.06 ± 0.1 with an ideal simulator.

**Table 3.** Results of beta regression for QNN’s predictive performance (F1 score) and configuration parameters.

|  | Breast Cancer Data |  | Synthetic Data |  |
| --- | --- | --- | --- | --- |
|  | Noisy Simulator | Ideal Simulator | Noisy Simulator | Ideal Simulator |
| Data Scaling Methods<sup>a</sup> |  |  |  |  |
| Standard | -0.22 (0.07)** | -0.21 (0.06)** | -0.29 (0.12)* | -0.22 (0.05)* |
| Min0-Max2Pi | 0.27 (0.07)** | 0.46 (0.07)** | -0.20 (0.13) | -0.31 (0.05)** |
| QNN Model Parameters |  |  |  |  |
| Ansatz Circuit - EfficientSU2<sup>b</sup> | 0.32 (0.06)** | 0.37 (0.06)** | 0.13 (0.11) | 0.29 (0.05)** |
| Ansatz Repetitions | 0.03 (0.01)* | 0.18 (0.01)** | -0.16 (0.02)** | 0.08 (0.01)** |
| Optimization Method - SPSA<sup>c</sup> | 0.45 (0.06)** | 0.21 (0.06)** | 0.60 (0.11)** | 0.16 (0.04)** |
| Optimization Level | 0.14 (0.02)** | 0.12 (0.02)* | 0.15 (0.04)** | 0.06 (0.02)* |
| Feature Map Repetitions | -0.13 (0.01)** | -0.02 (0.01)* | -0.44 (0.03)** | -0.06 (0.01)** |
| Input Gradients | 0.13 (0.06)* | 0.35 (0.06)** | -0.06 (0.11) | 0.09 (0.05) |
| Log-Likelihood | 1817.7 | 1932.8 | 1195.2 | 2308.8 |
| AIC | -3565 | -3796 | -2320 | -4548 |
| BIC | -3570 | -3600 | -2125 | -4353 |

<sup>a</sup> The baseline value is Min0-Max1.
<sup>b</sup> The baseline value is RealAmplitudes.
<sup>c</sup> The baseline value is COBYLA.
\* $p < 0.05$ (threshold)
\*\* $p < 0.01$

**Figure 3.**
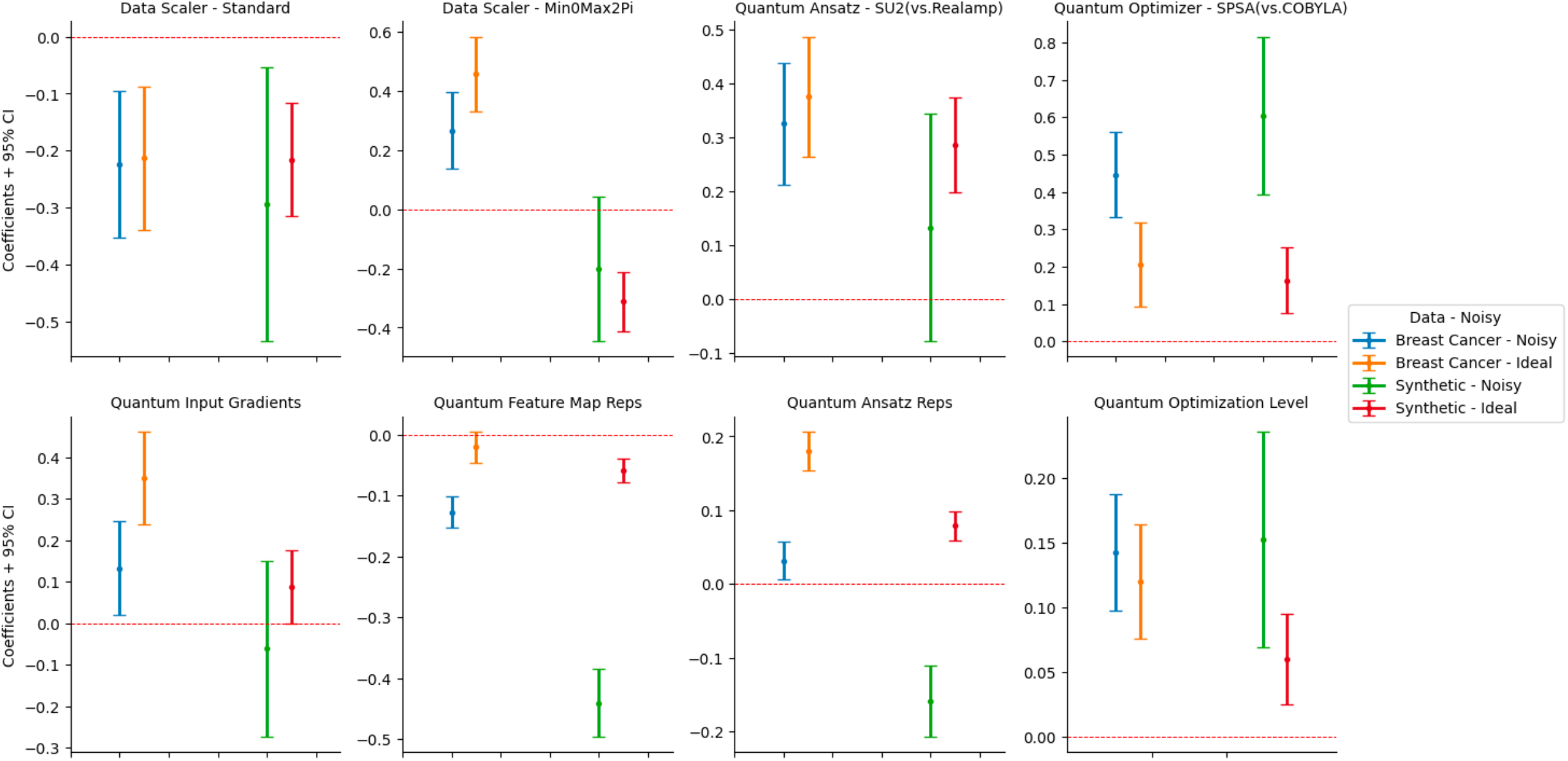
Beta regression results showing the impact of data scaling and configuration parameters on QNN performance by dataset and simulator type. All reported results represent coefficients of parameters regressed against the F1 score using the Beta regression method. The regression models included both individual effects and their first-order interactions.

In addition, increasing quantum transpiler’s level of circuit optimization improved the predictive performance of the QNN models. For the breast cancer dataset, the coefficients were 0.14 ± 0.02 using a noisy simulator and 0.12 ± 0.02 using an ideal simulator. For the synthetic dataset, the coefficient was 0.15 ± 0.04 using a noisy simulator and 0.06 ± 0.02 using an ideal simulator. Additionally, all QNN models performed better with the SPSA optimizer compared to the COBYLA optimizer. For the breast cancer dataset, the coefficient of the SPSA optimizer, compared to the COBYLA optimizer, was 0.45 ± 0.06 using a noisy simulator and 0.21 ± 0.06 using an ideal simulator. For the synthetic dataset, the corresponding coefficient was 0.60 ± 0.11 using a noisy simulator and 0.16 ± 0.05 using an ideal simulator.

Both using the EfficientSU2 ansatz circuit and increasing the circuit depth largely increased the predictive performance of QNN (Table 3). Compared to the RealAmplitudes circuit, applying the EfficientSU2 ansatz yielded higher F1 scores in three out of four cases across datasets and simulator configurations. For the breast cancer dataset using a noisy simulator, the coefficient of the Efficient SU2 ansatz, compared to RealAmplitudes, was 0.33 ± 0.06 and was 0.37 ± 0.06 using an ideal simulator. For the synthetic dataset using an ideal simulator, the corresponding coefficient was 0.29 ± 0.05 and using a noisy simulator, the coefficient was positive but insignificant. Similarly, the circuit depth, or the number of circuit repetitions, also increased F1 scores in the same three cases. For the breast cancer dataset using a noisy simulator, the coefficient was 0.06 ± 0.01 and using an ideal simulator was 0.18 ± 0.01. For the synthetic dataset using an ideal simulator, the corresponding coefficient was 0.08 ± 0.01.

Data scaling method and input gradients were two unique configuration parameters that had different impacts on the real-world dataset from the synthetic dataset. For the breast cancer dataset, scaling the data to a range of 0 and 2*p maximized the model’s predictive performance with both simulator configurations, 0.27 ± 0.07 with a noisy simulator and 0.46 ± 0.07 with an ideal simulator. For the synthetic data, scaling the data between 0 and 1, the baseline value, maximized the model performance. Also, while enabling input gradients had a positive association with F1 scores in the breast cancer dataset for both using a noisy simulator (0.13 ± 0.06) and using an ideal simulator (0.35 ± 0.06), this effect was not validated for the synthetic dataset.

### Weak interaction effects were observed between configuration parameters in QNN

We also explored potential first-order interactions and dependency of data scalers and model configuration parameters in our impact analysis (Supplementary Table 1). No common interaction effects were observed across all four QNN models (Fig. 4). Although small, several significant interaction effects were consistent throughout simulator configurations. Using a noisy simulator, the optimization method (optimizer) and input gradient showed a meaningful interaction effect. Enabling input gradients when using the SPSA optimizer reduced the model performance by −0.09 ± 0.03 or greater for both datasets.

**Figure 4.**
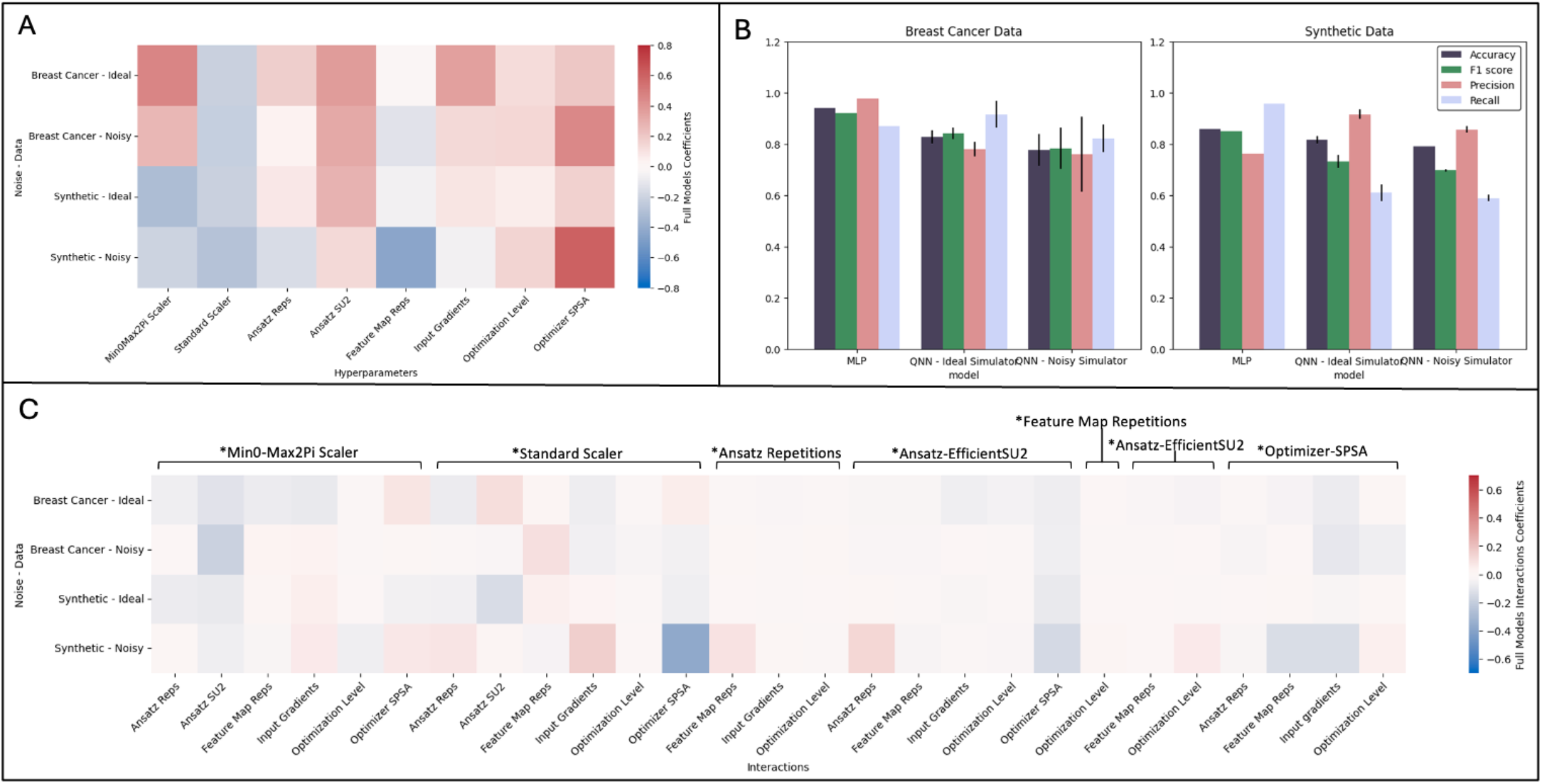
(A) Individual parameter impacts by data set and simulator type calculated from the beta regression results. (B) Comparison of the predictive performance of MLP and QNN across four performance metrics, accuracy, F1 score, precision, and recall. The best-performing parameter configuration for each QNN model was determined based on the average F1 score, and the mean values of the other three metrics were then calculated. MLP was modeled using the default hyperparameter settings in Scikit-Learn. (C) Interactions of data scalers and QNN model parameters by data set and simulator type calculated from the beta regression results.

Using an ideal simulator, the impact of circuit depth was affected by both input gradients and feature map repetitions. When using an ideal simulator and enabling input gradients, increasing the depth of circuit reduced F1 scores by at least −0.02 ± 0.01 for both datasets. Using an ideal simulator, increasing the depth of circuit mitigated the negative impacts of feature map repetitions (−0.01).

### QNNs Showed Prediction Performance Comparable to Classical NNs For the Synthetic

*Dataset*We compared the best-performing QNN models with the MLP model by dataset (Fig. 4). For the breast cancer dataset, the MLP outperformed the QNN models in all performance metrics but recall. While the MLP achieved an F1 score of 0.92, the QNN had F1 scores of 0.78 ± 0.08 using a noisy simulator and 0.84 ± 0.02 using an ideal simulator. Using an ideal simulator, the recall score was 0.92 ± 0.05, higher than using a noisy simulator (0.82 ± 0.05) and the MLP (0.87). For the synthetic dataset, both QNN models outperformed MLP in terms of precision. Using a noisy simulator, the mean precision score was 0.86 ± 0.01, whereas 0.91 ± 0.02 using an ideal simulator and 0.76 for the MLP model. The differences in performance between classical and quantum models were generally less marked for the breast cancer dataset.

## DISCUSSION

In this study, we demonstrated that the configuration of data scaling methods and model parameters can have a considerable impact on QNN model performance, paralleling prior literature on tuning classical machine learning models^20,26,27^. When properly optimized, our QNN architecture achieved performance comparable to that of an MLP, a result validated across two benchmark datasets.

Some configuration parameters consistently contributed to model performance across datasets and simulator configurations. Notably, using standardization for data scaling showed a negative effect, underlining the importance of proper feature scaling for both classical and quantum neural networks^15,28^. Effective data scaling is essential to the quantum encoding process where classical data is mapped onto quantum space^29,30^. One possible explanation for the suboptimal performance observed when using standardization for feature scaling in our study is the wide variation in feature ranges. Standardization is a scaling technique that normalizes data to have a mean of 0 and a standard deviation of 1. However, its sensitivity to outliers can distort the original meaning and variability of data and cause information loss, resulting in over- or under-representation of certain features and disrupt quantum encoding^28^. While further exploration is needed, our findings suggest considering alternatives to standardizing features, such as scaling to a fixed range using minimum and maximum values, when tuning a QNN model.

To encode our classical benchmark datasets, we used ZZFeatureMap. In this second-order Pauli-Z evolution circuit, each feature is encoded as a single-qubit rotation, and all qubits are fully entangled^31^. Literature has shown that increasing circuit depth and entanglement of a feature map can provide a quantum advantage when solving certain problems, such as those involving large feature spaces^32^. The underlying rationale is that greater circuit depth can enable the feature map to capture more complex data patterns. However, this approach must be balanced against data complexity, computational efficiency, and the capacity and compatibility of the available quantum resource^33^. Contrary to these expectations, our study found that increasing the number of feature map repetitions (i.e., circuit depth) consistently reduced model performance across all datasets, using noisy and ideal simulators. These mixed findings suggest that increasing the depth of a fully entangled feature map may introduce excessive computational complexity, thereby hindering the model’s ability to effectively learn the feature space and exacerbating potential decoherences with errors.

Although not unitary, some configuration parameters had notable impacts on model performance. The Efficient SU2 ansatz generally outperformed the RealAmplitudes circuit, except when training QNN on the breast cancer dataset with a noisy simulator. This finding also aligns with existing literature, which shows that the performance of ansatz architectures can vary depending on factors such as the dataset, the quantum computing system (e.g., cloud-based simulators or hardware), and the type of task^34,35^. For instance, one study comparing classical and quantum machine learning models for breast cancer detection using the same benchmark dataset (WDBC) observed no significant performance differences between the RealAmplitudes and Efficient SU2 ansatz circuits^34^. In contrast, another study on sentiment analysis reported that a QNN model using the EfficientSU2 ansatz outperformed one using RealAmplitudes when trained with 100 or 150 epochs, with all other parameters held constant^35^. Based on findings from both the literature and our study, we suggest that the EfficientSU2 ansatz may serve as a reasonable starting point for QNN model tuning.

Contrary to the negative impact observed when increasing the depth of feature map circuits, greater depth in ansatz circuits was generally associated with improved model performance. This positive effect was consistent across datasets and simulator configurations, except when training on the breast cancer dataset using a noisy simulator. Circuit depth influences a model’s capacity to represent complex data patterns by introducing additional layers of parameterized gates. While excessive depth can introduce issues such as overfitting or the barren plateau problem, where vanishing gradients hinder effective model training, findings from prior studies corroborate that increasing circuit depth can improve QNN performance, much like adding layers in classical neural networks^36–38^. While further validation is needed to generalize across factors such as problem complexity, circuit expressiveness, qubit entanglement, and hardware limitations, our findings, together with prior research, suggest that increasing the number of ansatz circuit repetitions may be a useful strategy for enhancing QNN model performance.

Transpilation is a process in circuit-based quantum computing that transforms an input circuit into a form compatible with the topology and hardware constraints of a specific quantum device or processor (backend). It plays a critical role as it compiles human-defined abstract gates into machine-compatible gates and optimizes the performance accordingly^39^. The transpilation workflow typically involves steps such as initialization, layout, routing, translation, and optimization of circuits prior to sending a QML model to the quantum backend and starting training (Fig. 1)^39^. Among these steps, optimization is essential to improve execution efficiency and improve circuit robustness against decoherence and errors to maximize the fidelity of the output circuit (Table 1, Fig. 1). The optimization level is a configuration parameter that controls the intensity of this effort, ranging from 0, no optimization, to 3, maximum optimization (Table 1). In our experiments, the optimization level was positively associated with QNN performance, and this effect was enhanced when using a noisy simulator (Table 2). While additional tuning and studies are needed to generalize this finding, initializing QNN parameter configuration with a transpiler optimization level above the median (for example, 2 in Qiskit) can be a practical starting point for QNN development. It is important to note, however, that higher optimization levels, while often requiring longer runtimes and more complex computing, do not guarantee an output of better quality due to the heuristic nature of transpiler optimization^40^

QNN performed better with the SPSA optimizer than with the COBYLA optimizer across datasets and simulator configurations. This finding is consistent with existing literature^41,42^. The choice of optimization method had a more pronounced effect using a noisy simulator. For the breast cancer dataset, the SPSA coefficient was 0.45 using a noisy simulator versus 0.21 using an ideal simulator (Table 2, Fig. 3). The difference was more pronounced when trained on the synthetic dataset, with SPSA yielding a coefficient of 0.60 using a noisy simulator 0.16 using an ideal simulator (Table 2, Fig. 3). These results align Pellow-Jarman, et al (2021)^41^, which attributed COBYLA’s reduced performance with a noisy simulator to its greater sensitivity to quantum noisy relative to SPSA. Taken together, SPSA can be a robust default choice for QNN model tuning, particularly in configurations where quantum noise is present.

In addition to evaluating individual effects, we also examined interaction effects between configuration parameters (Suppl. Table 1). One notable finding was the negative interaction between the optimization method and the use of input gradients when using a noisy simulator. QNN performed better when input gradients were not enabled when optimizing with SPSA and using a noisy simulator. A possible explanation is that SPSA, which relies on stochastic gradient approximations, may be more susceptible to challenges such as amplified noise and errors, vanishing gradients, and the barren plateau problem^4^. Input gradients, when enabled, may interact with quantum noise during parameter updates, further compounding these issues. In contrast, COBYLA is a gradient-free optimizer that employs linear approximations, making its performance less affected by the quantum environment.

While this study implemented QNN on a quantum simulator designed to closely imitate actual quantum hardware, simulators may not fully capture the complexities of real quantum environments. Future research should incorporate actual quantum processing units (QPUs) to better understand model performance in practical settings and assess the real-world applicability of QML. Expanding the scope to include a broader range of data types and structures, particularly in healthcare and biomedicine where data complexity and high dimensionality present unique challenges, will also be needed. A further limitation lies in the specific quantum algorithm and configuration parameters examined. As the field of QML rapidly evolves, introducing diverse algorithms with distinct configurations and model parameters, ongoing investigation into their impacts remains critical. Future studies should also seek to integrate advances in quantum computing technology with QML to increase the effectiveness and scalability of QML for real-world applications.

## CONCLUSION

In this study, we found that when pipeline configurations were optimized, QNN could achieve performance comparable to MLP, highlighting the critical role of configuration parameter tuning in quantum AI/ML development. Based on insights from the current study and existing literature, selecting data scalers with fixed ranges, limiting the number of feature map repetitions (n=1), increasing circuit depth (n>3), increasing transpiler optimization level (n>2), and the use of the SPSA optimizer may improve the performance of QNN in the IBM quantum architecture. This study contributes to the literature by evaluating the parameter choices in pipeline configurations and providing referential guidelines for QML model tuning. While current quantum computing limitations pose challenges, the potential of QNN provides a strong foundation for advancing quantum AI/ML research and its applications in healthcare and biomedicine.

## Data Availability

All data produced are available at: Street, W. Nick, William H. Wolberg, Olvi L. Mangasarian, Nuclear feature extraction for breast tumor diagnosis, Biomedical image processing and biomedical visualization, Vol. 1905, SPIE, July 1993, Pages 861-870, https://doi.org/10.1117/12.148698.

## Competing Interest Statement

W.L.S. was a technical consultant to HugoHealth, a personal health information platform (equity, fees); is a cofounder of Refactor Health, an AI-augmented data management platform for healthcare (equity); is a consultant for Detect, a point-of-care diagnostics company (equity, fees). B.N. is a consultant for Refactor Health, an AI-augmented data management platform for healthcare (equity). H.P.Y. is a consultant for Refactor Health, an AI-augmented data management platform for healthcare (fees). S.D. is now an employee at Royalty Pharma following the conclusion of study analysis.

## Funding

This work was partially supported by National Institutes of Health (NIH) grant number 1OT2OD032742-01.

---

**Supplementary Figure 1:**
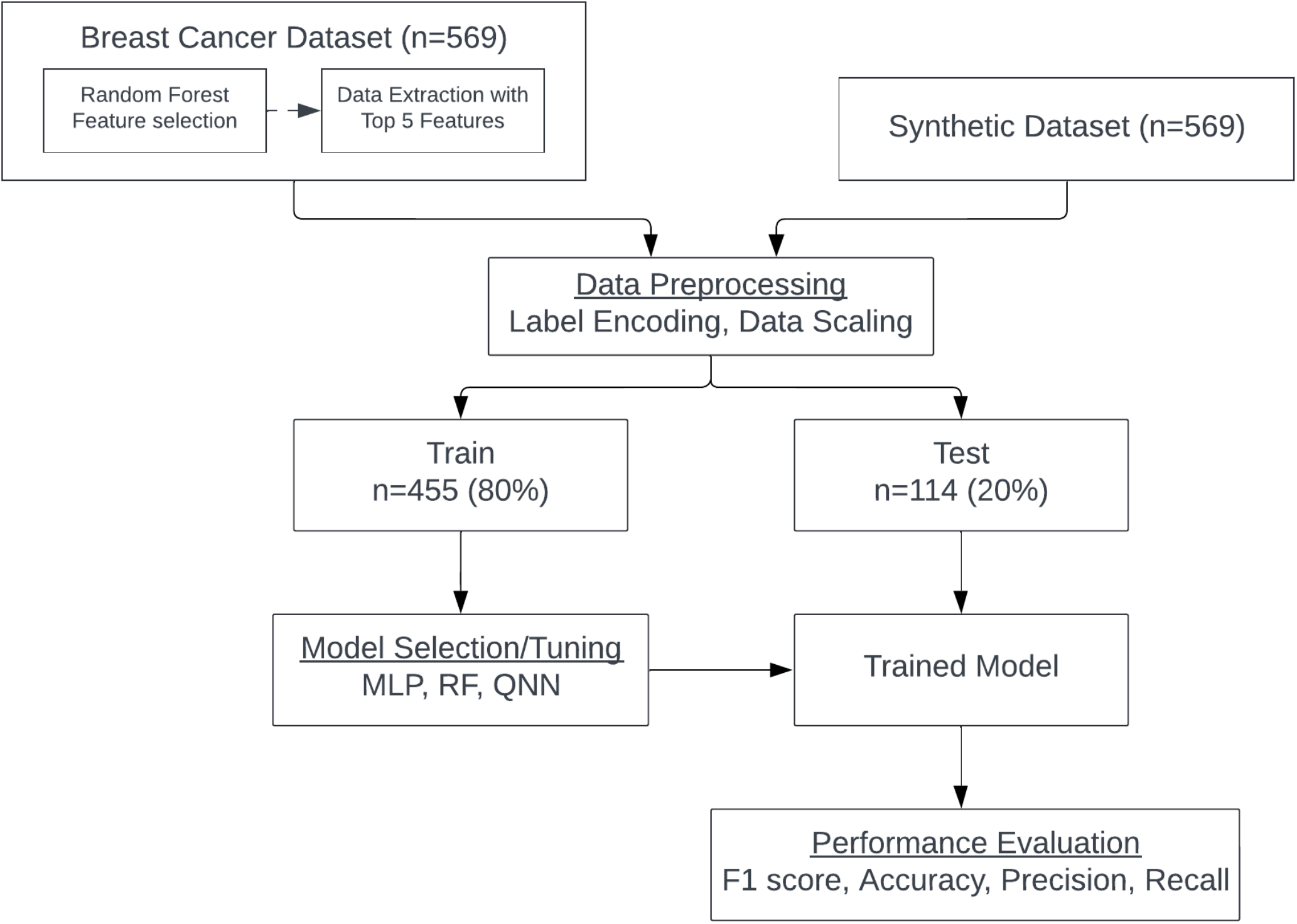
Study Design.

**Supplementary Table 1:**
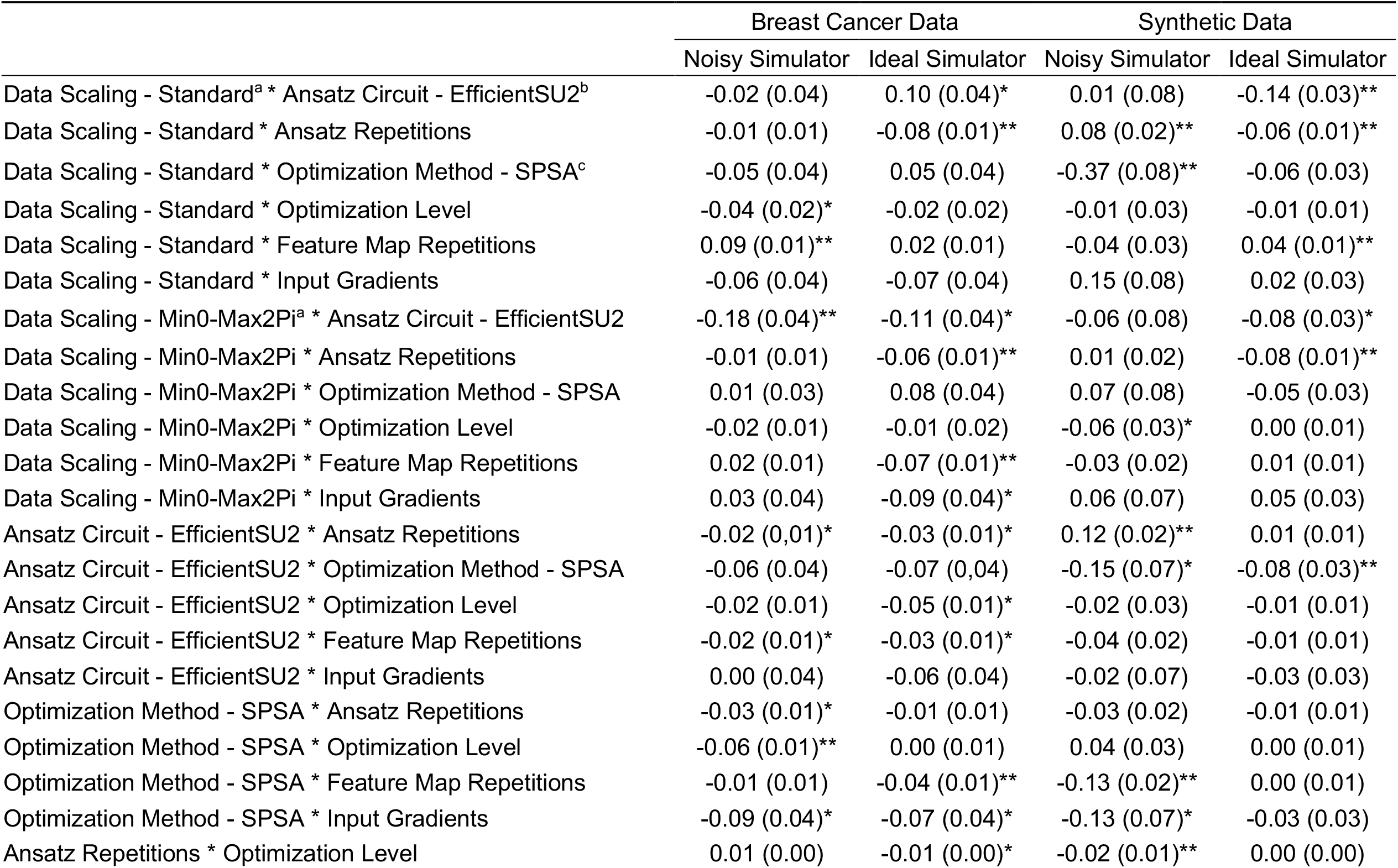
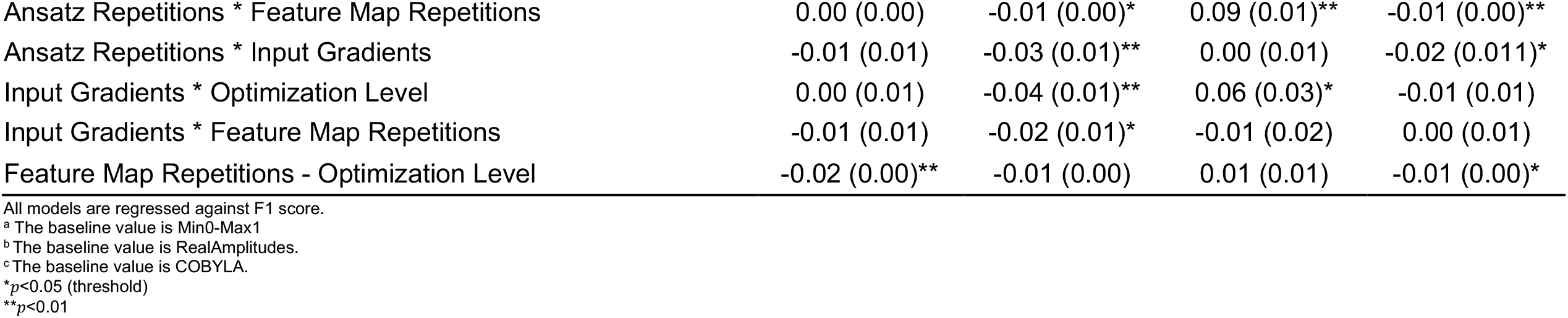
Interaction Effects in Beta Regression for QNN’s Predictive Performance (F1 Score) and Configuration Parameters.

## Notes

### Funding Statement

This study was partially supported by National Institutes of Health (NIH) grant number 1OT2OD032742-01.

### Author Declarations

The study only used data that were publicly available before the initiation of the study: https://archive.ics.uci.edu/dataset/17/breast+cancer+wisconsin+diagnostic

